# Genomic surveillance of SARS-CoV-2 tracks early interstate transmission of P.1 lineage and diversification within P.2 clade in Brazil

**DOI:** 10.1101/2021.03.21.21253418

**Authors:** Alessandra P Lamarca, Luiz G P de Almeida, Ronaldo da Silva Francisco, Lucymara Fassarella Agnez Lima, Kátia Castanho Scortecci, Vinícius Pietta Perez, Otavio J. Brustolini, Eduardo Sérgio Soares Sousa, Danielle Angst Secco, Angela Maria Guimarães Santos, George Rego Albuquerque, Ana Paula Melo Mariano, Bianca Mendes Maciel, Alexandra L Gerber, Ana Paula de C Guimarães, Paulo Ricardo Nascimento, Francisco Paulo Freire Neto, Sandra Rocha Gadelha, Luís Cristóvão Porto, Eloiza Helena Campana, Selma Maria Bezerra Jeronimo, Ana Tereza R Vasconcelos

## Abstract

The sharp increase of COVID-19 cases in late 2020 has made Brazil the new epicenter of the ongoing SARS-CoV-2 pandemic. Novel SARS-CoV-2 lineages P.1 and P.2, first identified respectively in Manaus and Rio de Janeiro, have been associated with potentially higher transmission rates and antibody neutralization escape. In this study, we performed a whole-genome sequencing of 185 samples isolated from three out of the five Brazilian regions, including Amazonas (North region), Rio Grande do Norte, Paraíba and Bahia (Northeast region), and Rio de Janeiro (Southeast region) aiming to identify SARS-CoV-2 mutations that could be involved in the surge of COVID19 cases in Brazil. Here, we showed a widespread dispersion of P.1 and P.2 across Brazilian regions. Except for Manaus, P.2 was the predominant lineage identified country-wise. P.2 lineage was estimated to have originated in February, 2020 and has diverged into new clades. Interstate transmission of P.2 was detected since March, but reached its peak in December, 2020 and January, 2021. Transmission of P.1 was also high in December. P.1 origin was inferred to have happened in August 2020. We also confirmed the presence of the variant under investigation (VUI) NP13L, recently described in the southernmost region of Brazil, to have spread across the Northeastern states. P.1, P.2 and NP13L are descended from the ancient B.1.1.28 strain, although during the first phase of the pandemic in Brazil presence of B.1.1.33 strain was also reported. We investigate here the possible occurrence of a new variant of interest descending from B.1.1.33 that also carries the E484K mutation. Indeed, the recurrent report of many novel SARS-CoV-2 genetic variants in Brazil could be due to the absence of effective control measures resulting in high SARS-CoV2 transmission rates. Altogether, our findings provided a landscape of the critical state of SARS-CoV-2 across Brazil and confirm the need to sustain continuous sequencing of the SARS-CoV-2 isolates worldwide in order to early identify novel variants of interest and to monitor for vaccine effectiveness.

## Introduction

One year after identifying the first case of SARS-CoV-2 infection in Brazil, the country is in a catastrophic situation with 11 million cases of COVID-19 and 265,000 deaths (https://coronavirus.jhu.edu/map.html). The initially dominant lineages B.1.1.28 and B.1.1.33 [1] have been gradually replaced by the new variant of concern P.1 and variant of interest P.2 harboring harmful mutations [2–4]. P.2 was firstly reported in November 2020 in samples from the state Rio de Janeiro and was estimated in previous works to have emerged in late July [3]. By December 2020, it was already prevalent in samples in Rio Grande do Sul, which borders the countries Uruguay and Argentina [5]. P.1, on the other hand, was first detected in the city of Manaus, north region of Brazil in early January 2021 [2,4], with a proposed emergence between April and mid/late December. Both lineages evolved within the B.1.1.28 clade carrying the E484K mutation in the receptor-binding domain (RBD) of the Spike protein. In addition to E484K, P.1 also harbors N501Y and K417T mutations in the RBD region, and both shared with the new variants of concern from the United Kingdom (B.1.1.17) and South Africa (B.1.351). Those three mutations are suggested to allow the viral escape from previous hosts’ immune responses [6–8]. This hypothesis is supported by the explosive spread of P.1 cases in Manaus and reports of reinfection involving both lineages.

During the first phase of the COVID-19 pandemic in Brazil, national long-distance travels from large urban centers in the Southeast region to North and Northeast states was attributed to be the source of the explosion of cases across the country [1]. Since mid-November, there has been an expected and dramatic surge in new COVID-19 cases, supposedly caused by a reduction of social distance levels due to end-of-the-year holidays and summer vacations (from December to February). This sharp increase in cases is attributed to the emergence of P.1 lineage, which has already been reported in several cities in Brazil [9,10]. Unfortunately, lineage pervasiveness and genomic diversity is still unknown or outdated in several Brazilian states. If the aforementioned mutations in P.1 and P.2 indeed promote escape from the host’s immune response, this information is crucial to elaborate measures to slow nationwide and worldwide spread. Furthermore, there is also a need to investigate whether there is a relationship between the variants and clinical outcomes, epidemiological patterns, response to vaccines and novel drugs.

Monitoring of P.1 lineage in Brazil is mostly executed using specific-targeted screening. Although this strategy is a valuable one to identify a chosen variants’ occurrence, it cannot correctly evaluate the relative frequency of this variant in the screened population. Furthermore, targeted sampling’s exclusive use prevents monitoring other lineages’ evolution. In this context, systematic random sequencing of SARS-CoV-2 samples was decisive, as our work shows, to evaluate the prevalence of P.1 and P.2 across Brazilian states and monitor the emergence of new variants of interest within known lineages. In order to reduce this information gap, we performed an epidemiological and genomic survey by sequencing 185 SARS-CoV-2 new genomes from three Brazilian regions, including states of Amazonas (North region), Rio Grande do Norte, Paraíba, Bahia (all three in the Northeast region) and Rio de Janeiro (Southeast region). Samples were originated from random RT-PCR positive results and obtained between December 2020 and February 2021.

## Materials and Methods

### Sample collection

In this work, a total of 185 participants from Amazonas (4), Rio Grande do Norte (44), Paraiba (43), Bahia (58), and Rio de Janeiro (36) states, representing the Brazilian North, Northeast, and Southeast regions, including 39 municipalities. Samples were selected from December 1st, 2020 through February 15th, 2021. Ninety-two males and 93 females were included, with age ranging between 11-90 years and with CT values between 8.70 to 29.00 (Appendix 1 Table S1). Samples from Manaus (Amazonas) were obtained from patients transferred to Paraíba in late January 2021. Samples from Rio de Janeiro were obtained from patients attending the Piquet Carneiro Polyclinic at the Rio de Janeiro State University (UERJ) to screen of COVID-19. Nasopharyngeal swabs were obtained from each participant and SARS-CoV-2 infection was diagnosed by RT-PCR using CDC/EUA protocol [11], OneStep/COVID-19 (IBMP, Brazil) AllplexTM 2019-nCoV (Seegene, South Korea) or nCoVqRT-PCR kits (Biomanguinhos, Fiocruz, Rio de Janeiro). The present study was approved by Ethical Review Board/Brazilian Commission of Ethical Study (Research Ethics Committee of: Universidade Federal Rio Grande do Norte - CAAE 36287120.2.0000.5537, CAAE 32049320.3.0000.5537, Universidade Federal da Paraíba - CAAE 30658920.4.3004.5183, Universidade Estadual do Rio de Janeiro - CAAE 30135320.0.0000.5259 and Universidade Estadual de Santa Cruz - CAAE 39142720.5.0000.5526). All data was analyzed anonymously.

### Next-generation sequencing and bioinformatics analysis

cDNA synthesis and viral whole-genome amplification were carried out following the Artic Network protocol (https://artic.network/ncov-2019). Amplicon libraries were prepared either using the Nextera DNA Flex kit (Illumina, USA). Sequencing was performed in a MiSeq System using MiSeq Reagent Kit v3 (Illumina, USA). Bioinformatic analysis was performed using an in-house pipeline for NGS data pre-processing, variant calling, and genome assembly as previously described [3,5,12]. The Wuhan-Hu-1 (NC_045512.2) sequence was used as the reference genome in our analysis.

### Phylogenetic analyses

The evolutionary position of the newly sequenced genomes was inferred using 1441 sequences from Brazil and 70 from other countries obtained from the GISAID database on February 25th, 2021. The Brazilian background sequences were selected following previous published protocol [13]. Modifications were clustering aligned sequences by 0.99985 similarity with CD-hit [14], keeping only the oldest record of each cluster and removing restrictions by country. Global sequences were added by selecting the oldest occurrence of each lineage in which the newly sequenced samples were classified into. Genome sequence from Wuhan-Hu-1 (NC_045512.2) sample was then added as an outgroup. All sequence alignment steps were conducted using MAFFT [15]. We used IQ-TREE2 [16] to infer the phylogeny of the final alignment. Simultaneously, the substitution model was selected with ModelFinder [16] using the global sequences as a proxy for genomic diversity within the larger alignment. Clade support was estimated using 10,000 replicates of bootstrap. To confirm the monophyly of NP13L clade and the new proposed lineage described in this work, we have also reconstructed their phylogenies with an expanded sampling to include all available sequences in GISAID that share their characteristic mutations.

We extracted P.1 and P.2 clades from the resulting maximum likelihood phylogeny to infer divergence dates and spatial dispersion with BEAST v1.10.4 [17]. After evaluating with TempEst [18] the correlation between root-to-tip distances and sampling dates (Figure S1), we selected the strict clock model to date P.1 divergence and the lognormal uncorrelated clock for P.2 [19]. Models used in both analyses were the Cauchy’s relaxed random walk for geographic coordinates [20,21] and the exponential growth coalescent tree prior. The MCMC was run through 10,000,000 steps with sampling every 10,000th and a burn in of 10% of the posterior results. We extracted ancestor coordinates using the SERAPHIM package [22] in R software.

## Results

The 185 newly sequenced genomes were assigned to 11 different lineages (Figure 1), with the majority belonging to P.1 (15.68%) and P.2 lineages (64.32%). Other lineages found were B.1.1.28 (4.86%), B.1.1.143 (4.33%), B.1.1.33 (4.33%), B.1.1.29 (2.70%), B.1 (1.62%), B.1.1.306 (0.54%), B.1.1.314 (0.54%), B.1.1.34 (0.54%) and B.1.212 (0.54%). The within-state relative lineages frequency revealed that P.2 was the most abundant lineage in Northeast and Southeast regions (Figure S2). Among the Northeast states, Rio Grande do Norte showed the highest occurrence of the P1 lineage (34.1% of the sequences obtained). Whereas, in the neighboring state of Paraíba, P.2 was the most frequent lineage (51.2% in this study) since late November 2020 [7], and P1 was only reported in early January 2021 (9.3%). Three lineages are described for the first time in the state (B.1.1.29, B.1.1.34 and B.1.212).

**Figure 1.**
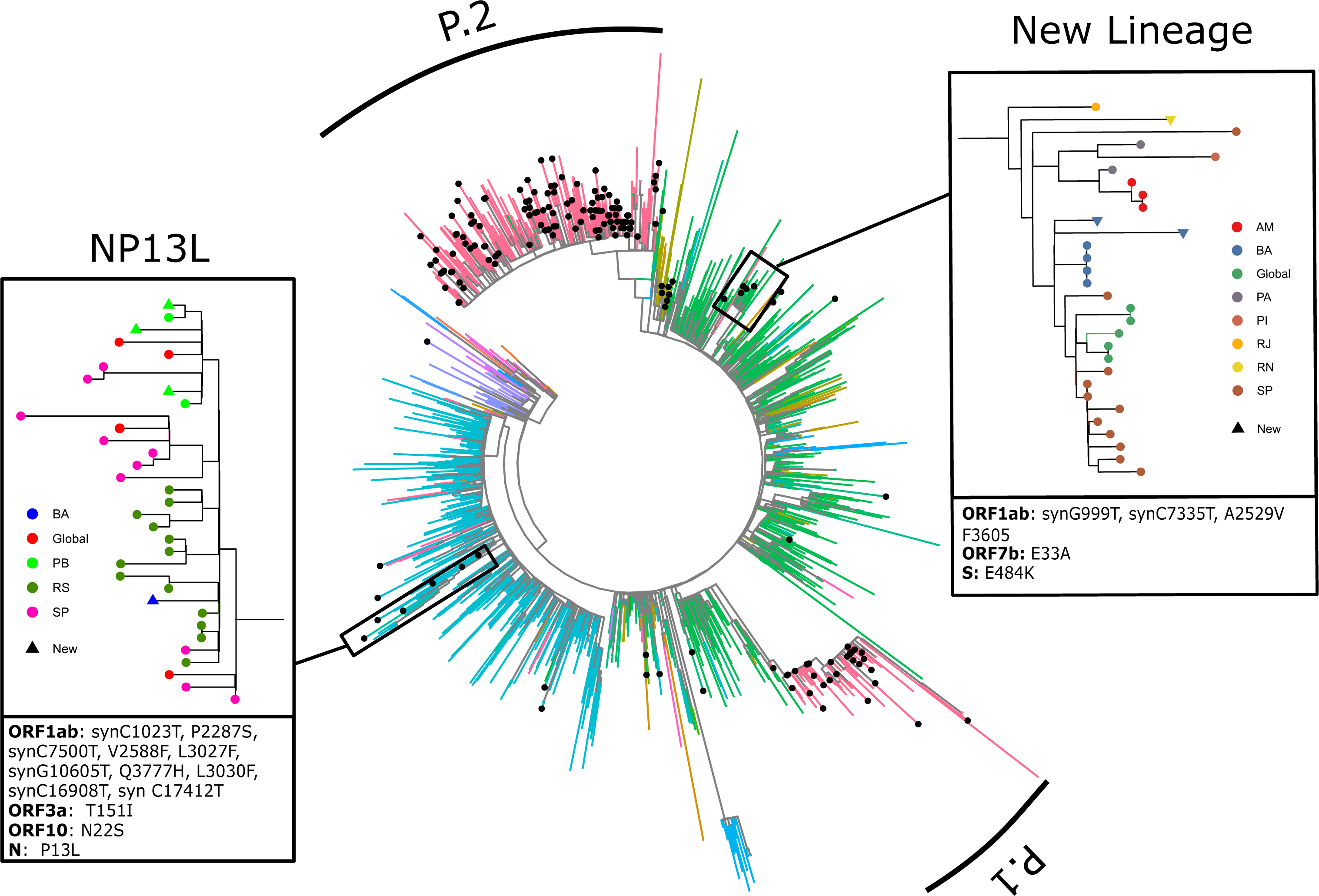
Phylogeny of 1696 SARS-CoV-2 genomes. Newly sequenced samples signalized by a point at the tip. Evolutionary lineages P.1 and P.2 are indicated by curved bars. In details, the new clade described in this work, originated from B.1.1.33, and the already proposed lineage NP13L. Colors in both smaller trees indicate the state of sample origin, and box below contains characteristic mutations of each lineage.

We identified 794 single-nucleotide variants (SNVs) across the 185 genomes sequenced, of which 49% were missense substitutions, 45% synonymous and 6% in non-coding regions of the genome (Figure S3). We also found three nonsense mutations in ORF8 (n = 2) and ORF7a (n = 1) in genomes from Rio de Janeiro, Rio Grande do Norte and Paraíba. We observed an elevated accumulation of mutation in the 3’UTR of the genome, mainly targeting ORF3 (subunits a, c and d), ORF9 (b and c), ORF8 and ORF7a (Appendix 1 Table 2). The nucleocapsid (N) protein and the subunit S1 of Spike protein showed the highest accumulation among the structural proteins of the SARS-CoV-2 genome. We found 16 SNVs targeting the receptor-binding domain (RBD) in S1, of which eight were missense variants, including K417T, N439K, L452R, S477R, E484K, N501Y, L518I, A522V.

A newly sequenced sample from Rio de Janeiro state was recovered as the first divergence within P.1 lineage (Figure 2A). Remarkably, this genome shows traces of intermediary evolution between B.1.1.28 and P.1, harboring 13 out of the 15 lineage-defining mutations according to Pango (https://cov-lineages.org/global_report_P.1.html). We did not observe two mutations (T20N, E92K) characteristic of P.1 clade. The evolutionary position of this new sample was confirmed by repeating the phylogenetic inference analysis with higher P.1 sampling and recovered the same results here described. This newly observed divergence pulls the estimated origin of P.1 lineage to mid-August 2020. Accordingly, interstate dispersal begins in September, leaving the state of Amazonas to northeastern states of Rio Grande do Norte and Paraíba (Figure 2B). The divergence between previously sequenced P.1 would happen in mid-October, giving rise to the most common variant. By November, the lineage was already widely distributed across the country with transmission originating in several states, including a reintroduction from Rio Grande do Norte to Amazonas. Interstate transmission reaches its peak in December, with new dispersion routes and maintenance of previous ones.

**Figure 2.**
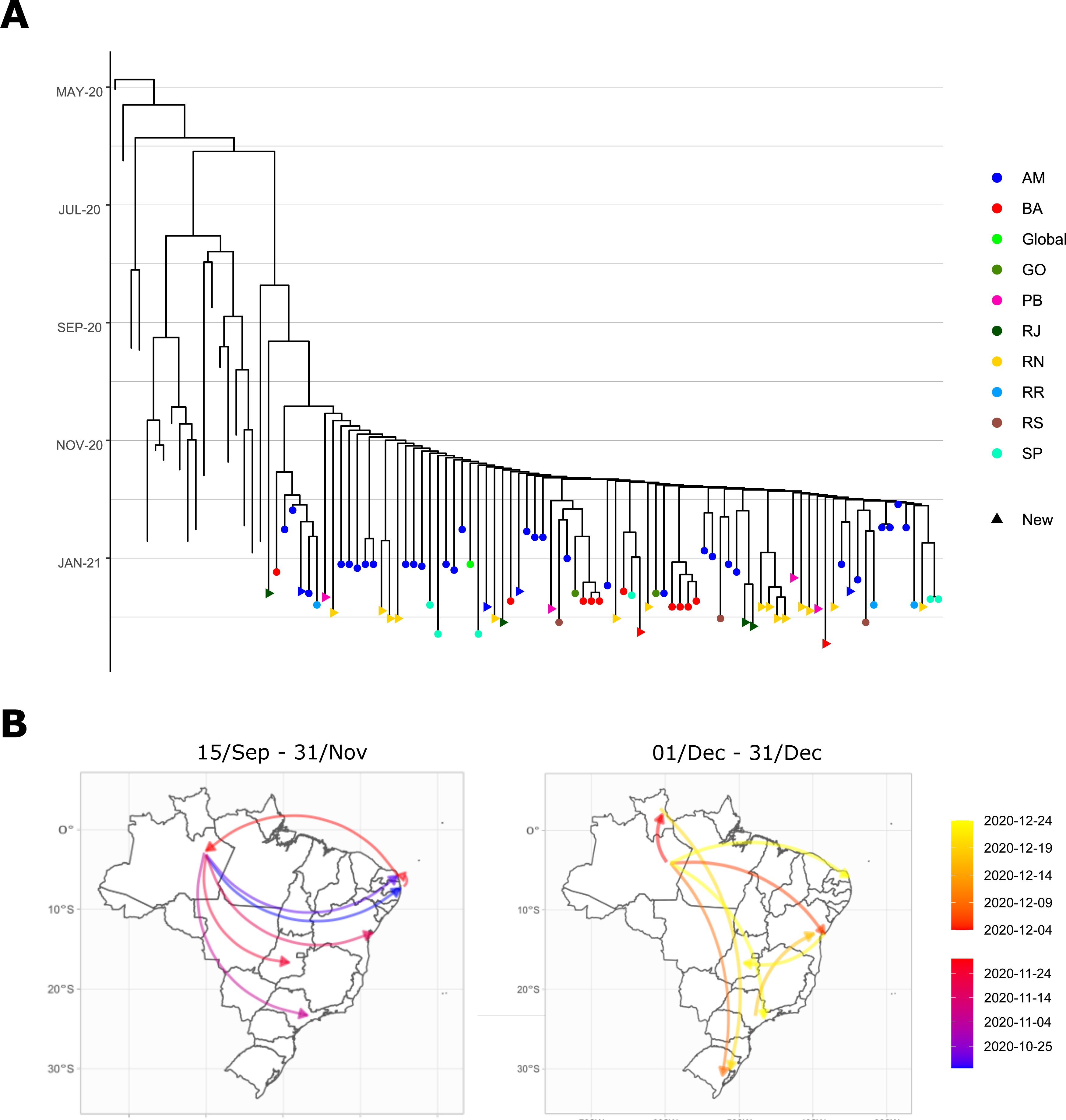
Divergence times within P.1 lineage (A) and its dispersion routes (B). Colors of tree tip points indicate the sample origin, while tips without a point represent the outgroup. Colors in the map indicate the date each interstate transmission route initiates.

While P.1 sequences are very similar due to their recency, we observed clear evolutive differentiation within P.2 lineage. The first within-clade diversification was estimated to have occurred in late February and the lineage went unreported until December [23], resulting in uncontrolled transmission across the country (Figure 2B). First introduction occurs simultaneously between São Paulo and Rio de Janeiro, followed by transmission to Brazil’s southernmost state of Rio Grande do Sul. From then onwards, a multitude of dispersion routes is observed between states. Mirroring P.1 behavior, interstate transmission of P.2 was also most intense during December and extended into January of 2021.

We have identified a monophyletic clade of 15 sequences within lineage B.1.1.33 that share a E484K variant. To confirm its monophyly, we have reconstructed this clade’s phylogeny while further increasing the sampling of B.1.1.33 sequences to contain all genomes with the E484K available at GISAID (Figure 1). All these extra samples fall within the described clade. Therefore, monophyly of the group was not disrupted by either the extensive B.1.1.33 outgroup sampling employed on the larger tree (100% bootstrap support) or by increasing the supposed ingroup. Also noteworthy, one of the newly-sequenced samples from Bahia of to this clade is the single B.1.1.306 reported in this work and additionally harbors a previously undescribed N501Y mutation in this lineage. We hypothesized that this new combination of mutations within B.1.1.33 might be due to Pango misclassification of this sample. Finally, we have also confirmed the monophyletic status (100% bootstrap support) of the proposed lineage NP13L [5], which emerged from B.1.1.28 in Brazil’s southernmost state and are now spread in the Northeast region. This result was, again, recovered even after increasing ingroup sampling with additional sequences available in GISAID. Finally, we report the occurrence of a single sample from the state of Rio Grande do Norte classified as B.1.1.29 that contains both E484K and N429K, uncharacteristic variants of the lineage.

## Discussion

The ongoing surge of SARS-CoV-2 in Brazil since the end of 2020 has turned the country into the epicenter of a very quickly spread of new variants [2,4]. In the present work, we have conducted a genomic surveillance of SARS-CoV-2 spread and evolution in historically undersampled regions of Brazil. Through the reconstruction of phylodynamics from P.1 and P.2 lineages, we have identified possible new variant lineages and past/current interstate transmission routes. We have also inferred the origin of P.1, suggested to be the lineage causing a drastic resurgence in COVID-19 cases [24], to have occurred around August. In contrast, phylogenetic analyses of P.2 indicate that the lineage originated in February 2020, when the virus was first reported in the country, and is evolving into differentiated clades.

Our genomic surveillance has evaluated the frequency of lineages currently circulating in each sampled state. As expected, proximity to the Amazonas state seems to be correlated to the pervasiveness of P.1 lineage, as exemplified by the variation observed in Rio Grande do Norte, Paraíba and Rio de Janeiro. The relatively low frequency of P.1 and high frequency of P.2 in our sample from the south of the state of Bahia, a region distant from large airports, may shed light on a much more complex relation between traveling and viral dynamics rather than guilt by association (i.e., mere vicinity). Indeed, previous works suggest that viral spread in smaller or distant cities may happen in a first-come-first-get dynamic, with one lineage overtaking the population [25–27]. Beyond south Bahia cities, this can be seen on Amazonas samples, where all four samples were from P.1 lineage. A significant advantage of low viral diversity is to decrease the likelihood recombination between lineages during a coinfection [5]. These results reinforce the importance of both local and international traveling restrictions as a preventive measure to slow the spread of the virus [28], measures still not enforced in Brazil and in many other countries. As an alternative, policies such as social distancing and early detection of more pathogenic variants could have curtailed the spread of P.1 and P.2 across states and unburdened the public health system [29–31].

Beyond inferred dispersal routes and spread, some lineages analyzed in this work require attention due to their evolutionary dynamics. First, we have observed that P.2 lineage has differentiated in several subclades between April and September of 2020 (Figure 3), all of them present in many states. The occurrence of P.2 subclades, in practice, means that epidemiological parameters, such as transmission rate, lethality, and immune response escape may vary within the lineage, hindering its containment [32]. If uncontrolled, the expected evolutionary course is for these subclades to evolve into whole lineages with exclusive mutations. Secondly, we have confirmed the monophyly of lineage NP13L [5], first described in January in Brazil’s southern regions. Since December, NP13L has been transmitted to Paraíba, and Bahia’s states, possibly from the Rio Grande do Sul. It has also been detected in England, Japan, and the Netherlands. Higher sampling and investigation of past and present transmission routes is urgent.

**Figure 3.**
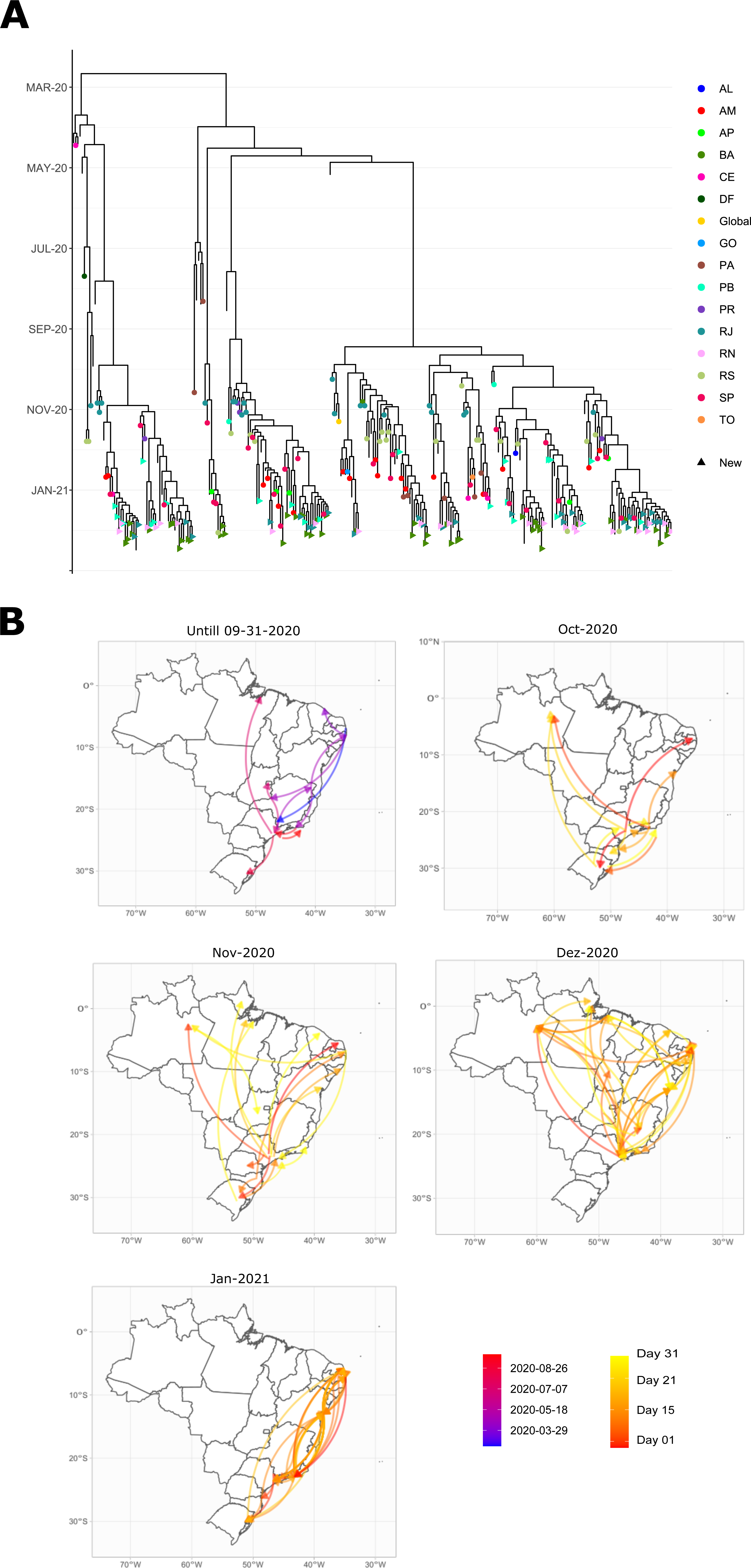
Divergence times within P.2 lineage (A) and its dispersion routes (B). Colors of tree tip points indicate the sample’s origin, while tips without a point represent the outgroup. Colors in the map indicate the date each interstate transmission route initiates.

Evidence of the occurrence of one possible new lineage derived from B.1.1.33 has also been demonstrated. According to our evolutionary inference, this lineage may have originated in Rio de Janeiro and disseminated across Brazil. This candidate new variant was already detected in samples from the United States, Ireland and Singapore. Remarkably, we observed the occurrence of E484K mutation in this new clade. This mutation was first detected in B.1.351 sequences from South Africa [33], but has now independently emerged in several lineages across globally, including P.1 and P.2 [2–4]. Another example of convergent evolution is the single sequence classified as B.1.1.306, which carries not only the mutation E484K inherited from the new lineage described here, but also the N501Y variant on the Spike protein gene. N501Y mutation was firstly identified in B.1.1.7 lineage in the United Kingdom [34] and recently detected in the P.1 lineage [2,10]. Finally, the third newly-detected convergent event described in this work is the E484K and N439K variants in a sample of B.1.1.29 from Rio Grande do Norte. The N439 mutation was also first detected at B.1.1.7 samples from the United Kingdom.

Convergent mutations seem to play an essential role in the evolutionary dynamics of SARS-CoV-2. Intense selective pressure from the immune system against prolonged infections may promote intrahost variants with higher adaptive value [12,35–39]. Previous studies have shown that both N501Y and E484K have independently emerged in patients with persistent infection [35,40]. Indeed, all convergent mutations aforementioned are somehow associated to viral escape from immune system response: N439K has shown to escape immune escape from both polyclonal and monoclonal antibodies [41,42]; E484K has been associated with escape from both vaccines and previous infections [8,23,43–45]; and N501Y leads to increased binding specificity to the receptor and is associated with high transmissibility while also escaping immune response [46,47]. Altogether, the combination of these mutations raises the variant’s fitness even higher, and increases the chance of the variant sequence becoming a new and dominant lineage [46]. Continuous monitoring of the convergent sequences here described is fundamental to follow their development and prevent spread in a worst-case-scenario.

Implementation of suitable genomic surveillance approaches through random sampling is a powerful tool to monitor known and new variants across the country. It can guide the elaboration of efficient governmental policies that avoid the collapse of the national healthcare system, as it is happening now. Both targeted screening and random sampling methods are complementary and congruent to an adequate evaluation of the current pandemic status. Of note, the analyses conducted here are highly dependent on broad sequence sampling through both time and space, which requires both technical and human resources training. Consequently, genomic surveillance is conducted only by a handful of laboratories, much less than needed to cover a continental-sized country as Brazil efficiently. Scientific collaborations such as conducted here bypasses regional barriers to monitor the advances of new and known lineages across states and foments an integrated analysis on the status of the pandemic in the country as a whole. Unfortunately, Brazil has become an open-air laboratory to the emergence and rapid dispersion of novel SARS-CoV-2 variants. Country-wide genomic surveillance is a significant step to better understand the origin and spread of new lineages.

## Data Availability

All sequence data is available in GISAID database. Accession numbers are located in Table S1.

## Figures

**Supplementary Figure 1. Correlation between root-to-tip distance and sequence sample dates**. Samples P.1 lineage (red) evolved under the same clock dynamics that outgroup sequences (black), whereas P.2 (blue) do not obey the strict clock model.

**Supplementary Figure 2. Frequency of SARS-CoV-2 lineages across Brazilian states**. Barplot showing the relative frequency of the 11 lineages found in this study in Amazonas (North region), Rio Grande do Norte, Paraíba, Bahia (all three in the Northeast region), and Rio de Janeiro (Southeast region).

**Supplementary Figure 3. Genomic characterization of SARS-CoV-2 mutations identified**. Distribution of single-nucleotide variants (SNVs) found in the 185 genomes sequenced in this study. Each vertical line represents the relative variant frequency in the total number of genomes sequenced and its target protein products. The receptor-binding domain (RBD) highlighted in red showed the main mutations associated with the variant of concern P.1 and variant of interest P.2. Density plot shows the accumulation of mutations across the SARS-CoV-2 genome.

## Tables

**Table S1. Sample information**.

**Table S2. Frequency of mutations in SARS-CoV-2 genome.**.

**Table S3. Acknowledge to GISAID samples**.

## Acknowledgements

We would like to thank all authors and the administrators of the GISAID database, allowing this genomic epidemiology study to be properly conducted. A full list of acknowledgment is available in Table S3. A list acknowledging those from different institutions that participated in this study follows bellow:

## Workgroup Members

### LABIMOL/ENDEMIAS/UFPB

Álisson Emannuel Franco Alves, Ana Beatriz Rodrigues dos Santos, Brena Ferreira dos Santos, Bruno Henrique Andrade Galvão, Daniela Letícia Torres da Silva, Fabio Marcel da Silva Santos, Fabrine Felipe Hilário, Gabriel Rodrigues Martins de Freitas, Marília Gabriela dos Santos Cavalcanti, Mayara Karla dos Santos Nunes, Moises Dantas Cartaxo de Abreu Pereira, Naiara Naiana Dejani, Sandrelli Meridiana de Fátima Ramos dos Santos Medeiros, Sergio Dias da Costa Junior, Talita Nayara Bezerra Lins, Wallace Felipe Blohem Pessoa

### LAFEM/UESC

Mylene de Melo Silva, Renato Fontana, Renata Santiago Alberto Carlos, Galileu Barbosa Costa, Hllytchaikra Ferraz Fehlberg, Amanda Teixeira Sampaio Lopes, íris Terezinha Santos de Santana, Fabrício Barbosa Ferreira, Luciano Cardoso Santos, Luane Etienne Barreto, Pérola Rodrigues dos Santos, Laíne Lopes Silva de Jesus, Thiago Silva Gonçalves, Gabriela Andrade Coelho Dias

### POLICLÍNICA PIQUET CARNEIRO/UERJ

Kennedy Martins Kirk, Claudia Henrique da Costa, Rogerio Rufino, Claudia Henrique da Costa, Renata Salles Miraldi Oliveira, Gabriella da Silva Alves, Allan Motta Leal Pontes, Sandra Pereira C. Vilas Boas, Vania Maria Almeida de Souza, Vinícius Miranda Porto, Jeane de Souza Nogueira

### DACT/CCS/CB/IMT/UFRN

Igor de Farias Domingos, Antonia Cláudia J. Câmara, Ivanise M. Moretti Rebecchi, Ana Cláudia G. Freire, Marcela A. Galvão Ururahy, Vivian N. Silbiger, André D. Luchessi, Antonnyo P. D. Lima, Thiala S. J. da Silva Parente, Carlos R. do Nascimento Brito, Kátia Castanho Scortecci, Susana M. Gomes Moreira, Daniella R. A. Martins Salha, Leonardo C. Ferreira, Waleska R. D. B. de Medeiros, Dayse Santos Arimateia, Arthur R. de Araújo Oliveira, Fernanda M. de Azevedo, João F. Rodrigues Neto, Glória Regina de Gois Monteiro, Paulo Ricardo P. do Nascimento, Ingryd Camara Morais, Francisco Paulo Freire Neto, Eliana L. Tomaz Nascimento, Iara Marques Medeiros, Ana Rafaela de Souza Timóteo

### LNCC/MCTI

Luciane Prioli Ciapina, Rangel Celso Souza, Éllen dos Santos Correa, Guilherme Cordenonsi da Fonseca, Vinícius Prata Klôh, Eduardo Wagner

## Funding statement

This work was developed in the frameworks of Corona-ômica-RJ (FAPERJ = E-26/210.179/2020 http://www.faperj.br/) and Rede Corona-ômica BR MCTI/FINEP (FINEP = 01.20.0029.000462/20 http://www.finep.gov.br/; CNPq = 404096/2020-4 https://www.gov.br/cnpq/pt-br). A.T.R.V is supported by Conselho Nacional de Desenvolvimento Científico e Tecnológico - CNPq (303170/2017-4) and FAPERJ (E-26/202.903/20). R.S.F.J is a recipient of a graduate fellowship from CNPq. A.P.L is granted a post-doctoral scholarship (DTI-A) from CNPq. SMBJ was supported by Ministério da Educação https://www.gov.br/mec/pt-br (UFRN Covid Task Force), Conselho Nacional de Desenvolvimento Científico e Tecnológico (440893/2016-0) and by JBS https://jbs.com.br/. E.S.S.S was supported by Fundação de Amparo à Pesquisa do Estado da Paraíba – FAPESQ http://fapesq.rpp.br/ (003/2020) and Laboratórios de Campanha MCTI/FINEP (0494/20 - 01.20.0026.00). L.F.A.L was supported by Conselho Nacional de Desenvolvimento Científico e Tecnológico and Coordenação de Aperfeiçoamento de Pessoal de Nível Superior https://www.gov.br/capes/pt-br. The funders had no role in study design, data collection and analysis, decision to publish, or preparation of the manuscript.

## Notes

### Competing Interest Statement

The authors have declared no competing interest.

### Funding Statement

This work was developed in the frameworks of Coronaomica RJ (FAPERJ = E-26/210.179/2020) and Rede Coronaomica BR MCTI/FINEP (FINEP = 01.20.0029.000462/20, CNPq = 404096/2020.4). A.T.R.V is supported by Conselho Nacional de Desenvolvimento Cientifico e Tecnologico CNPq (303170/2017.4) and FAPERJ (E-26/202.903/20). R.S.F.J is a recipient of a graduate fellowship from CNPq. A.P.L is granted a post-doctoral scholarship (DTI.A) from CNPq. SMBJ was supported by Ministerio da Educacao (UFRN Covid Task Force), Conselho Nacional de Desenvolvimento Cientifico e Tecnologico (440893/2016.0) and by JBS. E.S.S.S was supported by Fundacao de Amparo a Pesquisa do Estado da Paraiba FAPESQ (003/2020) and Laboratorios de Campanha MCTI/FINEP (0494/20 01.20.0026.00). L.F.A.L was supported by Conselho Nacional de Desenvolvimento Cientifico e Tecnologico and Coordenacao de Aperfeicoamento de Pessoal de Nivel Superior.

### Author Declarations

The present study was approved by Ethical Review Board/Brazilian Commission of Ethical Study (Research Ethics Committee of: Universidade Federal Rio Grande do Norte - CAAE 36287120.2.0000.5537, CAAE 32049320.3.0000.5537, Universidade Federal da Paraiba - CAAE 30658920.4.3004.5183, Universidade Estadual do Rio de Janeiro - CAAE 30135320.0.0000.5259 and Universidade Estadual de Santa Cruz - CAAE 39142720.5.0000.5526). All data was analyzed anonymously.

### Summary of Updates

Correction of figures order.

