## Supplementary figures and images for "Genomic surveillance of SARS-CoV-2 tracks early interstate transmission of P.1 lineage and diversification within P.2 clade in Brazil"

### Supplementary Figure 1

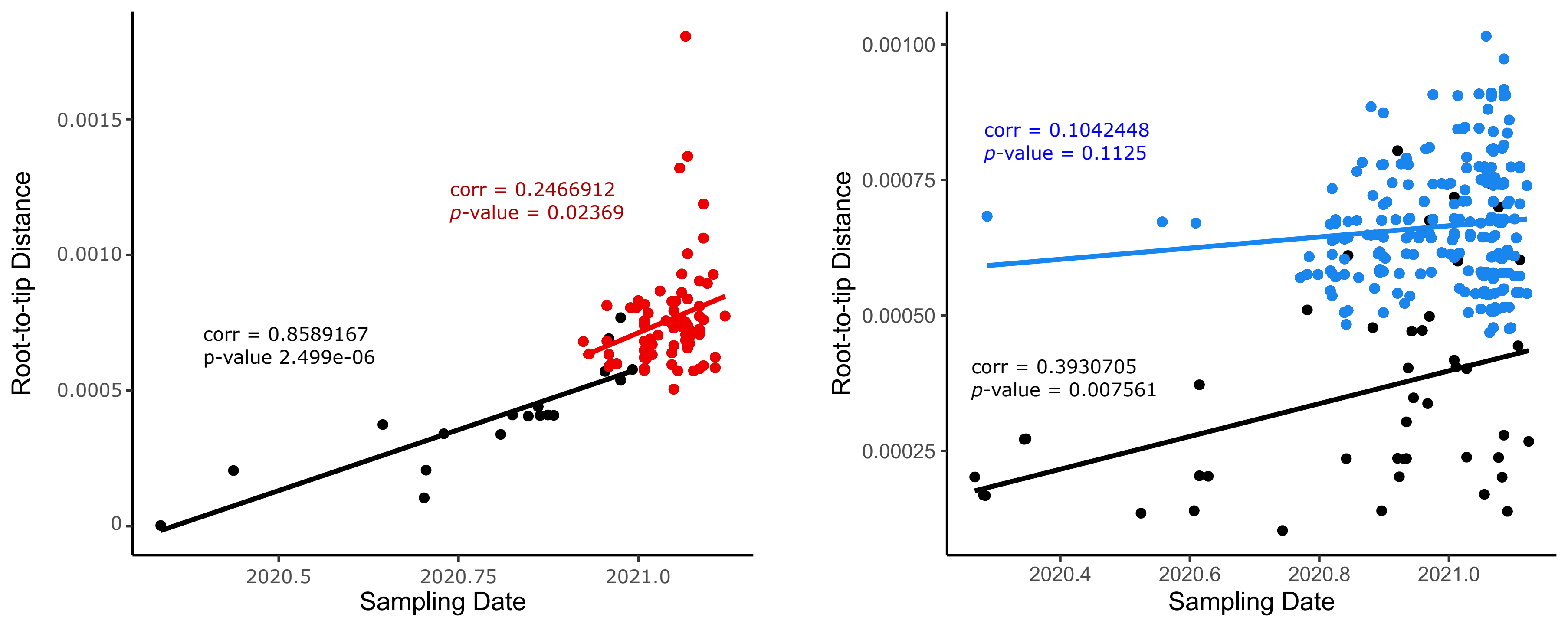

### Supplementary Figure 2

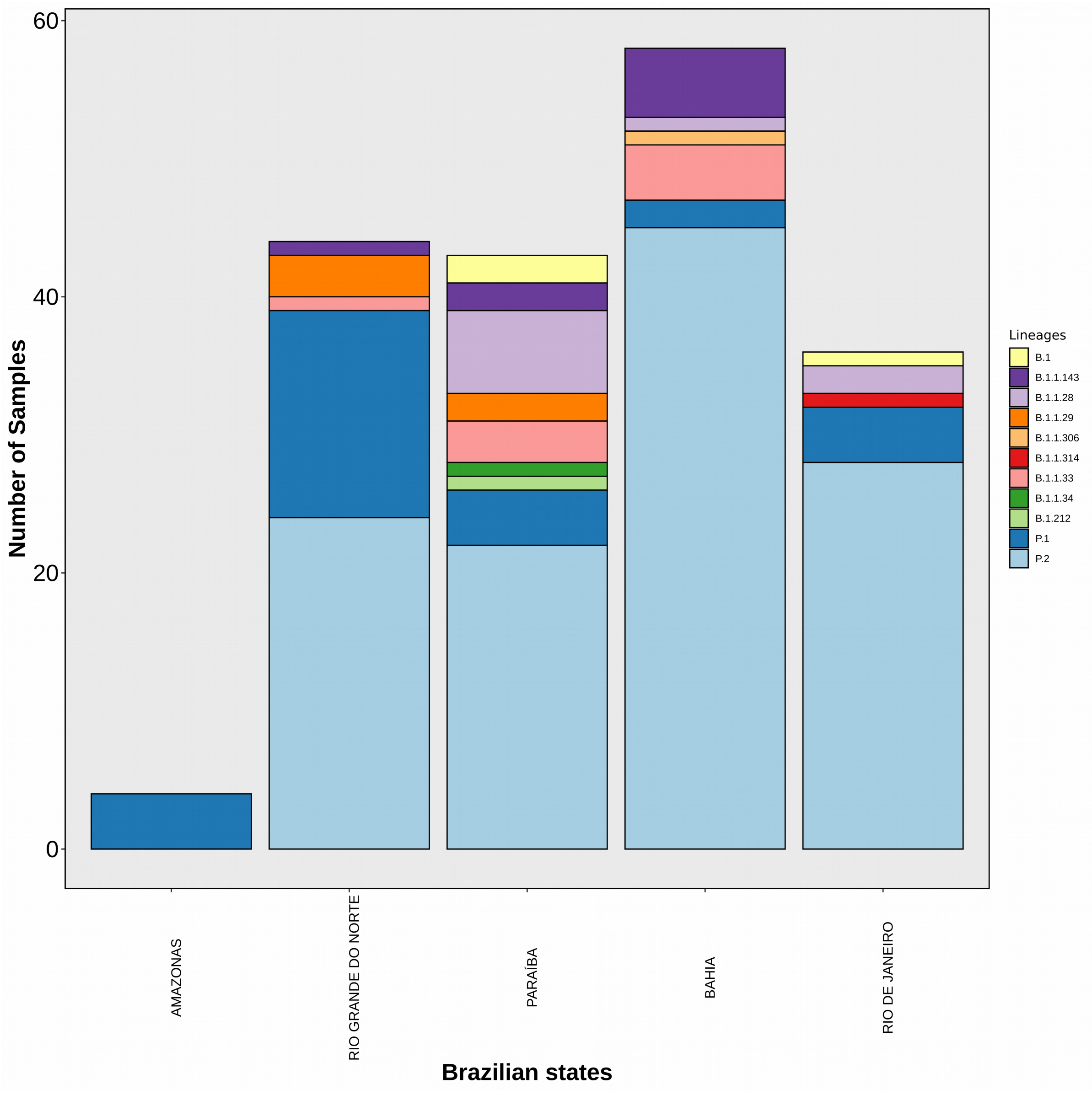

### Supplementary Figure 3

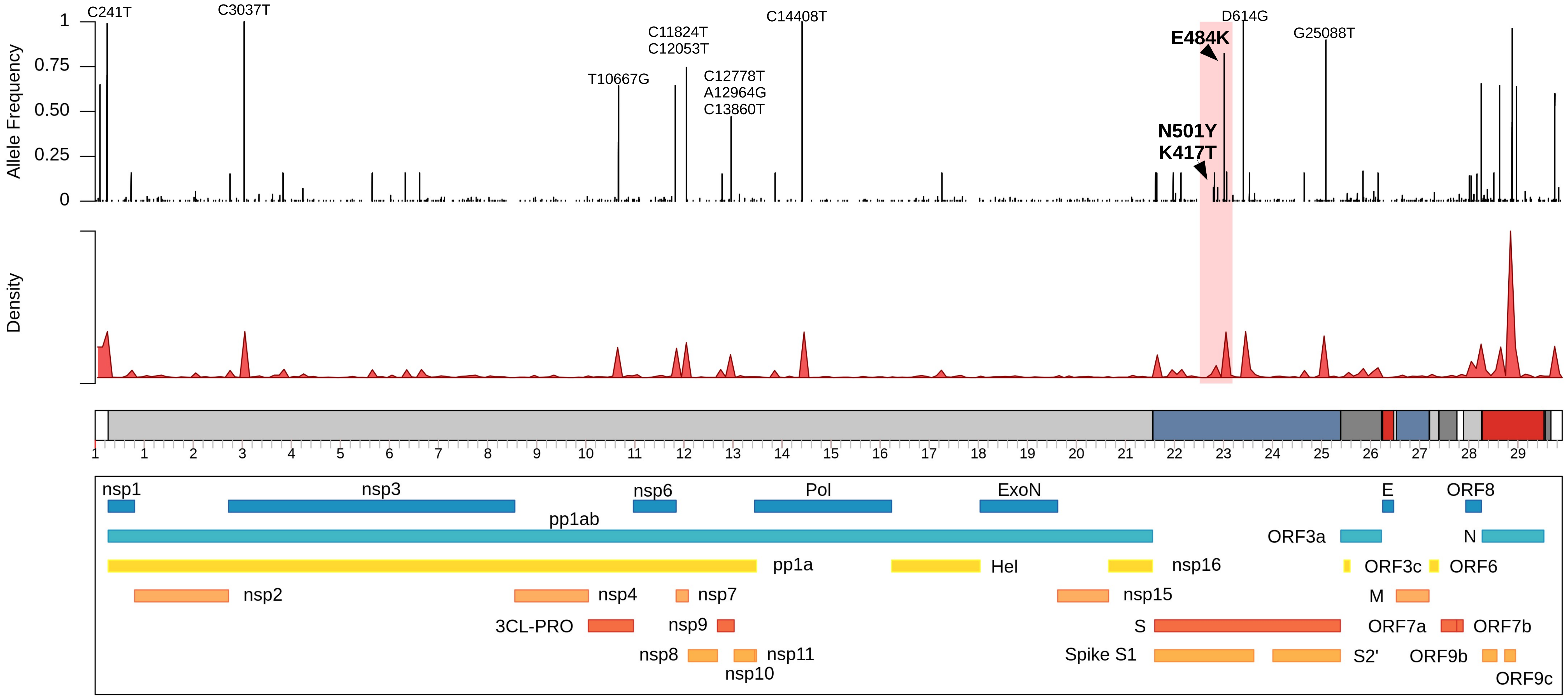
